# ‘Anatomy of SARS-CoV-2 outbreak of ‘vaccinated’: An observational case-control study of COVID-19 breakthrough infections, COVID-19 appropriate behavior and anti-spike-IgG response as a correlate of protection in Medical college students at Rural Medical College, India

**DOI:** 10.1101/2022.01.27.22269902

**Authors:** Monika Chavan, Sowmya Gayatri, Suvarna Patil, Janhavi Deshpande, Arvind Yadav, Prasanna Nakate, Yogendra Shelke, Anup N Nillawar

**Author notes:** **(Corresponding Author)** Dr Anup N Nillawar, Postal Address: Same as Above, Email Id. Email id. Authors’ Contribution: The first authors; MC and SG contributed equally. MC and SG oversee the investigations and data collection and contributed in manuscript writing, JD and SP provided crucial clinical data about hospitalized patients, AY reviewed manuscript and contributed in data collection, PN and YC gave RT-PCR inputs, AN conceptualized the paper, prepared study design and prepared CRF and did manuscript writing and oversee laboratory investigations.

## Abstract

**Introduction:** Breakthrough infections about severe acute respiratory syndrome coronavirus 2 (SARS-CoV-2) have been reported worldwide in both partially or completely vaccinated individuals irrespective of the type of vaccine. India, also emerged as the vaccine powerhouse producer, initiating the world’s one of the largest vaccination drives since January 16, 2021, with two vaccines named, BBV-152(COVAXIN™) and AZD1222 (COVISHIELD™). Breakthrough cases are reported from all over the globe for all kinds of vaccines. This is the investigation report into the outbreak of breakthrough COVID-19 infections at one of the medical colleges in Rural India unraveled in early October 2021. This report underscores the role of COVID-19 appropriate behavior along with vaccination and the role of IgG in evaluating immunity generated through vaccination.

**Objective:** The study objective was to 1. Clinically characterize the recent breakthrough infection of COVID-19 (SARS-CoV-2) infection among (students) vaccinated in BKL Walawalkar Rural Medical College. 2. To evaluate ‘COVID-19 appropriate behavior’ (CAB) in cases and controls. 3. To evaluate COVID-19 anti spike IgG in the cases in comparison with the control

**Methods:** A total of 74 students studying at BKL Walawalkar Rural Medical College and vaccinated for COVID-19 were included in the study. RT-PCR diagnosis was done from 5 to 10 October 2021. The breakthrough infection in the cases was characterized using self-assessment questionnaires in comparison to the controls. The cases were assessed clinically and also using biochemical parameters. Both cases and controls were also assessed for their adherence to COVID-19 appropriate behavior using a separate semi-quantitative questionnaire and scoring system.

**Results:** In our study, out of the total subjects, 50% of Covaxin recipients had experienced vaccine breakthrough infection and 20% of Covishield recipients experienced breakthrough infection. Also, 6 out of the 35 cases were asymptomatic, and the rest were either having mild symptoms. None of them required any hospitalization or O2 therapy. The CAB score was lower in the cases when compared to controls. All the vaccine recipients show seroconversion. Anti-spike IgG antibodies titers are dynamic over time and across the clinically distinct groups. Irrespective of varying IgG titer, the vaccine protects against severity and possibly mortality.

**Conclusion:** The need for better awareness of COVID-19 appropriate methods as an alternative to and additive to vaccination is necessary to control the transmission of COVID-19 infection and decrease the disease severity. Vaccines are effective against preventing severe disease and possibly mortality. The use of serum anti-spike COVID-19 IgG is restricted to know the status of SARS-CoV-2 seroconversion.

## INTRODUCTION

The vaccination campaign for COVID-19 in India was started on January 16, 2021, using two vaccines; AZD1222-ChAdOx1-S (Covishield)(Non-replicating adenovirus vector based), manufactured in India by Serum Institute of India through a license from AstraZeneca-Oxford.^1^ and BBV152 (Covaxin)(Whole virus inactivated Type), an indigenous vaccine developed by Bharat Biotech in collaboration with the Indian Council of Medical Research (ICMR).^2^ From May 1^st^ onwards vaccination is open to all individuals (>18 yr of age) in India. The efficacy after administration of two doses of the vaccines irrespective of the interval between the doses has been reported as 63.1% and 78% for AZD1222 (ChAdOx1-S) and BBV152 respectively.^1,3^ Vaccines are widely regarded as the most effective means of halting and terminating the COVID-19-19 outbreak. However, due to a variety of factors vaccine efficacy in the real world may differ from that reported in clinical trials. Because no vaccine provides 100 % protection against the disease, and newer virus types periodically evolve mechanisms to evade the vaccine-induced antibody response, a small percentage of people will contract COVID-19-19 despite being fully vaccinated. Also, presumably, other preventive measures such as COVID-19 appropriate behavior which determines the degree of exposure to the virus, could play a major role in preventing breakthrough infections.^4^ Further, strict adherence to such behavior would also allay the severity of infection. Breakthrough infections about COVID-19-19, refer to the incidence of SARS-CoV-2 infections in individuals who have already been partially or completely vaccinated with any authorized COVID-19-19 vaccine.^5^ Many population based studies have documented the incidence of breakthrough infections as high as 10-15%.^4,6^ According to the comprehensive Nation-wide study undertaken by ICMR in between 5 March to 3 June 2021, showed surge in cases in India was mainly due to delta variant B.1.617.2.^7^ The current study attempts to characterize COVID-19 breakthrough infections to concerning clinical symptomatology, vaccination status, and inclination and awareness towards COVID-19 appropriate behavior. This is pursued by using a self-assessment semi-quantitative questionnaire in simple understandable language. This study would provide crucial evidence in understanding not only the vaccine effectiveness but also the need for strict adherence to appropriate behavior in preventing transmission and decrease in the severity of the disease.

### OBJECTIVES

1. To clinically characterize recent COVID-19 breakthrough infection of vaccinated subjects in BKL Walawalkar Rural Medical College during 5 to 10 October 2021.
2. To evaluate ‘COVID-19 appropriate behavior’ (CAB) in cases and controls.
3. To evaluate COVID-19 anti spike IgG in the cases in comparison with the control

## MATERIAL AND METHODS

### Study background

BKL Walawalkar rural medical college and hostels are the closed knit campus situated in the rural Konkan region where college and hostel campus is situated away from village dwellings and with very little possible contact with the outsiders and even with the hospital premises. In such a campus, in the last week of September 2021, we found one index case having mild upper respiratory tract infection (URTI) and came COVID-19 positive on RT-PCR. This triggered a thought to check for all the students if they are having any spread as they live in closely placed hostel rooms and closed sitting arrangements in offline teaching classes. Boys and girls have separate hostel buildings and are situated far apart. All the students were vaccinated against SARS-CoV-2 almost at the same time period, around April 2021.

### The rationale for selecting the study group

The study group is in many ways a closed group. Being in the same age group and staying in the hostel campus before and after vaccination and during this investigation. Hence, there is less chance of bias on the exposure to external sources of infection. Hence, there is a considerable chance that the transmission was from the index case and later on the spread to the rest of the class and was also restricted only to the close contacts of the infected. In a way, this can be described as a point source outbreak. Also, all of them were vaccinated at around a similar time. (March-April 2021) This helps in understanding the clinical effectiveness of the vaccine. To assess the kinetics of antibody response, we evaluated titers of COVID-19 anti-spike IgG.

## LABORATORY TESTING

All nasopharyngeal swab samples were tested in an automated PCR machine, Roche LC-96. This targets two genes that are ORF-1 and E gene. Anti-spike IgG antibodies against SARS-CoV-2 were quantitatively estimated in an automated chemiluminescence electro assay (CLIA) - based platform, ARCHITECT i1000SR (Abbott diagnostics) using a commercial quantitative kit SARS-CoV-2 IgG LJ quant. The cut-off value is 50 AU/ml.

## STUDY SUBJECTS

A total of 74 students studying at BKL Walawalkar rural medical college, Dervan, Maharashtra, India, who were all vaccinated for COVID-19 19 infection in March and April 2021(first dose of vaccine) were included in the study. All the students who tested positive for COVID-19 19 by RT-PCR with/without clinical symptomatology and at least one dose of COVID-19 vaccination were selected as cases. Similarly, all the RT-PCR negative tested students with no clinical symptomatology and with a history of at least one dose of vaccination were selected as controls. For finding out the best matching controls, we briefed the cases about the study design and asked them to choose the best matching control based on the contact time between them and amongst their peers. The study included 35 cases and 39 controls. Case record forms (CRF) were prepared to gather the following information: 1. Demographics, History of previous COVID-19 19 infection, and vaccination details (As per the government of India policy, two doses of either BBV152 (Covaxin) or AZD1222 (Covishield) vaccine at least 4 weeks apart were available for administration since January 2021 with no mixing of doses allowed) 2. COVID-19 clinical symptomatology, assessed by a semi-quantitative ‘self-assessment clinical questionnaire’ and a self-assessment questionnaire on ‘COVID-19 appropriate behavior (CAB)’ before and after this outbreak (CRF as Annexure). Both the questionnaires were prepared in an easily understandable manner and consisted of questions, each with a score from 0 to 10. The self-assessment questionnaire to assess ‘COVID-19 appropriate behavior (CAB) included five topics (a. wearing of mask b. Use of hand sanitizer c. frequent hand washing d following social distancing e. following travel restrictions) with each topic having a maximum score of 10. The total maximum score possible in the clinical questionnaire is 230 and for CAB is 50. (Annexure attached). The study protocol was approved by the Institutional Ethics Committee. Written consent was obtained from the subjects before the initiation of the study.

## STATISTICAL ANALYSIS

Continuous data has been presented as median (25th-75th centiles) while categorical data has been presented as percentages. Spearman correlation was used to analyze the association between IgG titers and clinical scores. The data is not normally distributed the Mann Whitney U test was used to compare two groups. Statistical analysis was performed using Statistical Package for the Social Sciences version 21.0 (SPSS, Inc.; Chicago, IL, USA) for Windows. p-Value <0.05 was considered statistically significant.

## RESULTS

A total of 74 students studying at BKL Walawalkar rural medical college, were included in the study. Table 1 shows the basic characteristics of the study subjects. Among the breakthrough cases, 32 had received 2 vaccination doses (fully vaccinated according to vaccination schedule) while, 3 had received one dose of vaccine. Among the 39 matched controls, 33 had received 2 and 6 had received one vaccination dose respectively Out of the whole cohort, 6 had received AZD1222/Covishield while 68 had received BBV152/Covaxin. (Table no. 2).

**Table 1:**
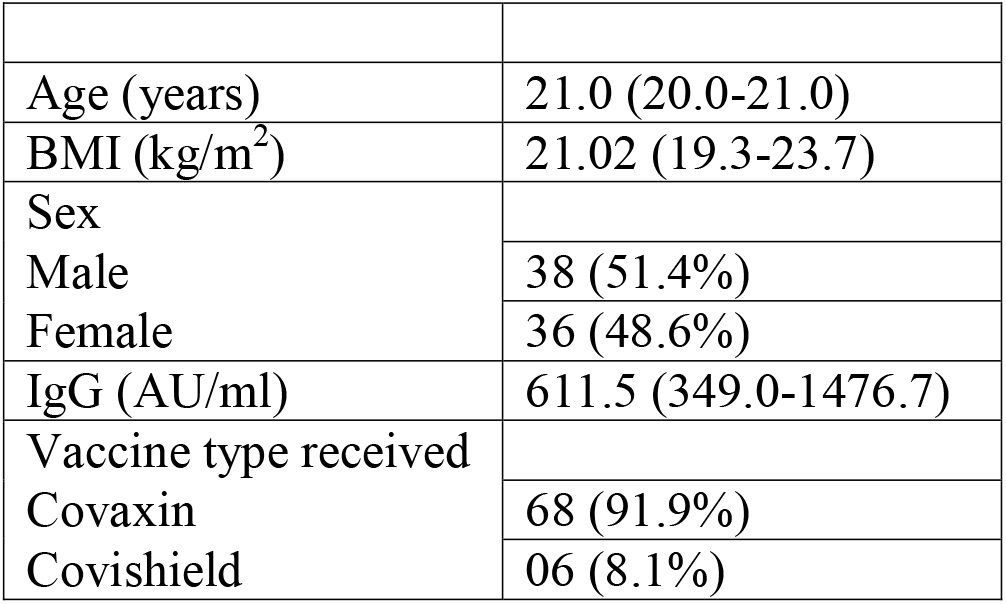
Basic Characteristics of Cohort:

|  |  |
| --- | --- |
| Age (years) | 21.0 (20.0-21.0) |
| BMI (kg/m<sup>2</sup>) | 21.02 (19.3-23.7) |
| Sex |  |
| Male | 38 (51.4%) |
| Female | 36 (48.6%) |
| IgG (AU/ml) | 611.5 (349.0-1476.7) |
| Vaccine type received |  |
| Covaxin | 68 (91.9%) |
| Covishield | 06 (8.1%) |

**Table 2:** Demographic and Clinical Characteristics of breakthrough cases receiving Covaxin and Covishield.

| Variable | Covaxin recipients<br>(n=68 ) |  | Significance | Covishield recipients<br>(n= 6 ) |  | Significance |
| --- | --- | --- | --- | --- | --- | --- |
|  | Case<br>(n=34) | Control<br>(n=34) |  | Case<br>(n=1) | Control<br>(n=5) |  |
| **Age(median,IQR)** | **20** | **21** | **0.000** | **19** | **21** | **NA** |
| **Sex:** |  |  |  |  |  |  |
| **Male** | **15** | **19** |  | **1** | **3** |  |
| **Female** | **19** | **15** |  | **0** | **2** |  |
| **BMI** | **22.0** | **20.1** | **0.059** | **23.4** | **22.2** | **NA** |
| **Symptomatic:** |  |  |  |  |  |  |
| **(Yes)** | **28** | **NA** |  |  |  |  |
| **If symptomatic, clinical Score on the scale of 230(mean)** | **33.9** | **NA** |  | **15** | **0** |  |
| **Asymptomatic** | **6** | **NA** |  | **0** | **0** |  |
| **Mean duration between second dose and Infection** | **150** | **88** |  |  |  |  |
| **Antibody positive against S protein** |  |  |  |  |  |  |
| **IgG titer(mean)** | **2993.5** | **444.5** | **p=0.036** | **3093** | **2095.6** | **NA** |
| **COVID-19 appropriate behavior(On scale of 50)** | **36** | **43** | **p=0.000** | **44** | **39.6** | **NA** |

(Case Arm: At least 1 dose of COVID-19 vaccine+ RT-PCR positive breakthrough COVID-19 infection)
(Control Arm: Age and ecology matching group with At least 1 dose of COVID-19 vaccine+ RT-PCR Negative)

1. Demography: The median age was 21.0 years. The median BMI of the study subjects was 21.02 kg/m^2^. The median Concentration of IgG was 726.0 (AU/ml). The mean antibody IgG titer between cases and control (irrespective of vaccine type) were 2996.3 and 656.2 respectively. (p=0.048). Additionally, we noted higher BMI in cases. (p=0.05) (Table No 2).
2. 50% of Covaxin recipients had experienced vaccine breakthrough infection and 20% of Covishield recipients experienced breakthrough infection. The average duration between the second dose and confirmed SARS-CoV-2 infection by RT-PCR was 150 days; in Covaxin recipients and 88 days; in Covishield recipients.
3. Clinical Charctecteristics: 6 out of 35 cases were asymptomatic, 29 positive cases with minimal symptomatology, 47% had anosmia and 41 % had a loss of taste. The most common symptom was runny nose and fever found in 50 % of patients. None had lower respiratory signs; none of them required hospitalizations or Oxygen support. All the symptoms were restricted to the upper respiratory tract. Out of 35, 10 were investigated for routine blood inflammatory markers and none had CRP>20 mg/L. Only 2 cases had CRP at 10 mg/L. None had increased D-Dimer above normal levels. The clinical score (calculated out of 23 symptoms, each symptom gauged on the scale of 0 to 10, on self-assessment questionnaire) is directly correlated with IgG titers (r=0.589, p=0.000). Subtly, this tells IgG titer increases with aggravation in clinical symptomatology.
4. COVID-19 Appropriate Behavior (CAB) Score: (Score was calculated on 5 criterion, each carrying 10 points, maximum score possible 50). And to our surprise, the mean CAB scores for cases and control were 36.2 and 42.5 respectively. (p=0.000). Both cases and controls improved on their CAB score post-outbreak.
5. Serum IgG titer: IgG antibodies against SARS-CoV-2 were quantitatively estimated in an automated chemiluminescence electro assay (CLIA) - based platform, ARCHITECT i1000SR (Abbott diagnostics). The present cohort (case; group2 and control; group3) was investigated for IgG after an average of 150 days of vaccination. Group 1 was vaccinated individuals with IgG measured at an average of 60 days of vaccination. Additionally, we compared IgG titers with a group (Group 4) having moderate to severe SARS-CoV-2, hospitalized, and needing oxygen support. (Table 3)
  a. All the vaccine recipients (Covaxin and Covishield recipients) showed seropositivity and antibody titer >50AU/ml. (Kit Manufactures’ cut-off).
  b. In the control group of the present study (Group2, Table 5), mean IgG levels are = 656 AU/ml. This analysis is done after the average lapse of 150 days. IgG levels are significantly lower when done after a wider time-lapse and it indicates waning antibody titers. Even though the titers are done in 2 different groups, they are enough indicating about waning antibodies.
  c. Table 4 indicates, additional infection with SARS-CoV2 bolsters IgG titers in vaccinated individuals.
  d. As the secondary objective was to know the SARS-CoV-2 anti-spike IgG titer and its association with immunity to SARS-CoV-2 re-infection. As shown in table 3, IgG titer varies across the groups, likewise; in ‘only vaccinated group’ (Group1), Group with mild breakthrough SARS-CoV-2 infection (Group 3), And patients with moderate to severe breakthrough infection (Group 4), who needed hospitalization and various levels of Oxygen support. Though the number in group 4 is less and no controlled matching was possible with the rest of the groups, the fact which remains assuring is that none of the patients with severe breakthrough infections also died. This trend needs to be ascertained and reinforced with more data. This table showed, though the IgG levels are varying across the groups and they are showing a waning trend, vaccination is helping to prevent severe disease and mortality. (Group 3 and 4 in Table 4)

**Table 3:**
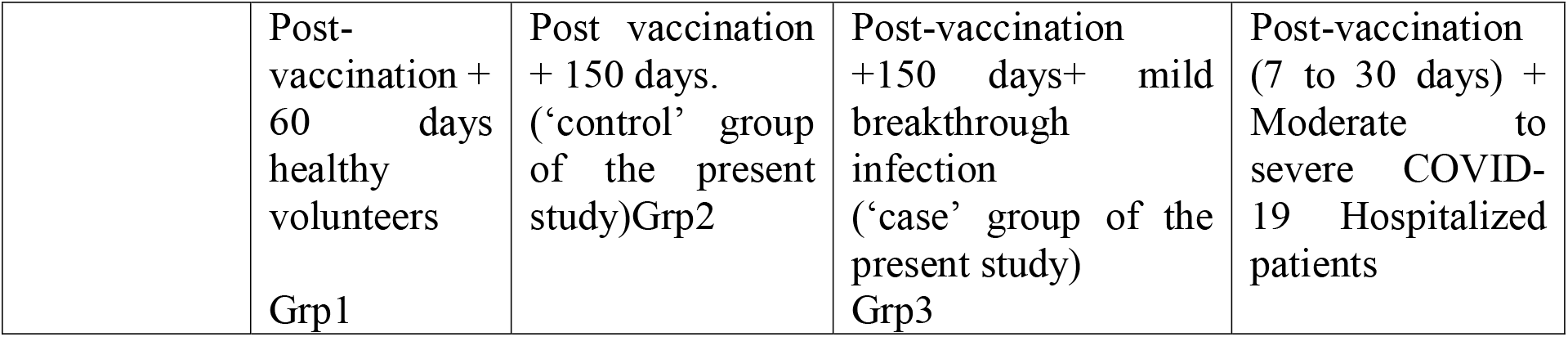
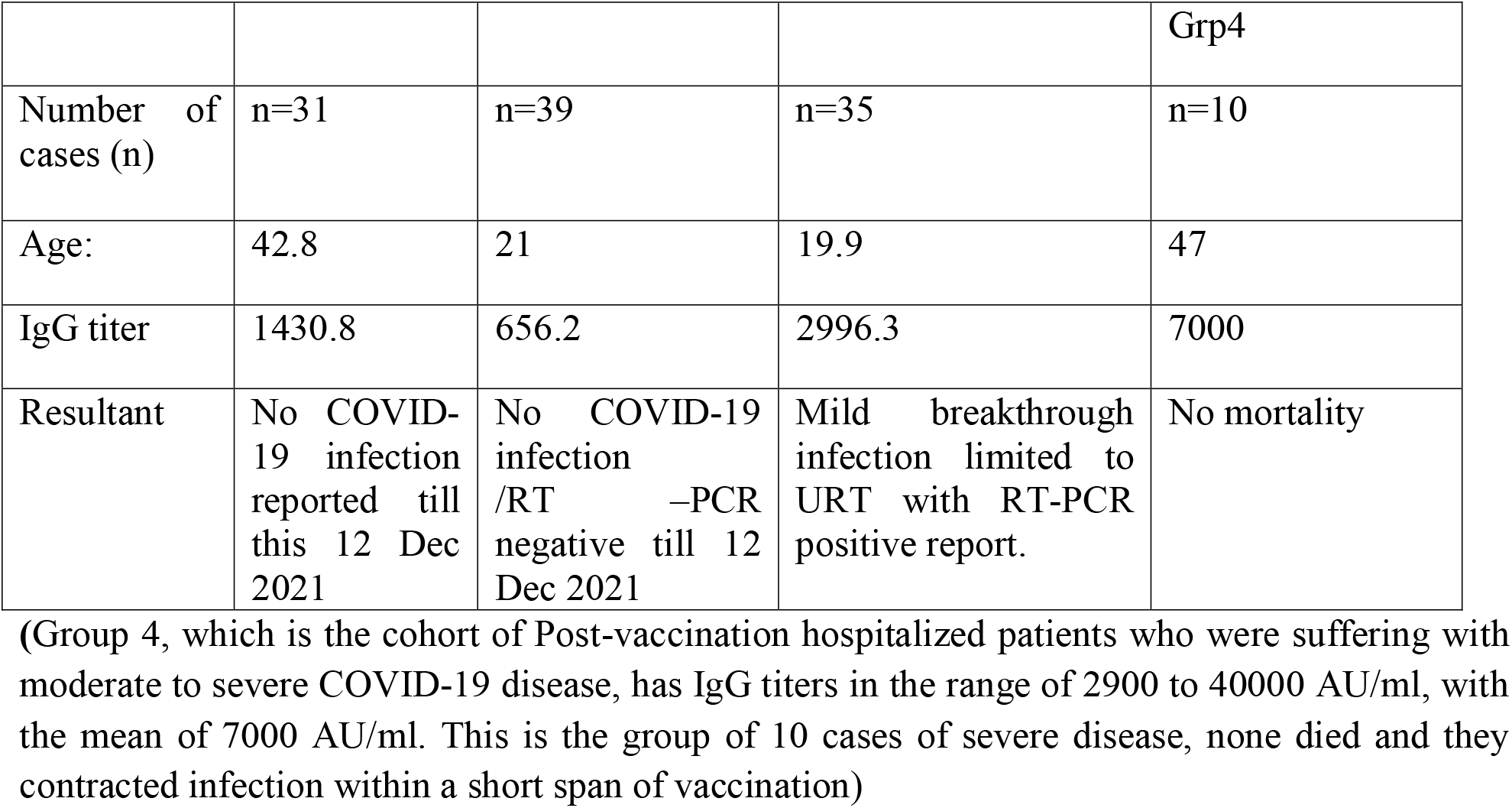
Comparison of IgG titers with additional data sets:

|  |  |  |  |  |
| --- | --- | --- | --- | --- |
|  | Post-vaccination + 60 days healthy volunteers<br>Grp1 | Post vaccination + 150 days. ('control' group of the present study)Grp2 | Post-vaccination +150 days+ mild breakthrough infection ('case' group of the present study)<br>Grp3 | Post-vaccination (7 to 30 days) + Moderate to severe COVID-19 Hospitalized patients |
|  |  |  |  | Grp4 |
| Number of cases (n) | n=31 | n=39 | n=35 | n=10 |
| Age: | 42.8 | 21 | 19.9 | 47 |
| IgG titer | 1430.8 | 656.2 | 2996.3 | 7000 |
| Resultant | No COVID-19 infection reported till this 12 Dec 2021 | No COVID-19 infection /RT –PCR negative till 12 Dec 2021 | Mild breakthrough infection limited to URT with RT-PCR positive report. | No mortality |

(Group 4, which is the cohort of Post-vaccination hospitalized patients who were suffering with moderate to severe COVID-19 disease, has IgG titers in the range of 2900 to 40000 AU/ml, with the mean of 7000 AU/ml. This is the group of 10 cases of severe disease, none died and they contracted infection within a short span of vaccination)

**Table 4:**
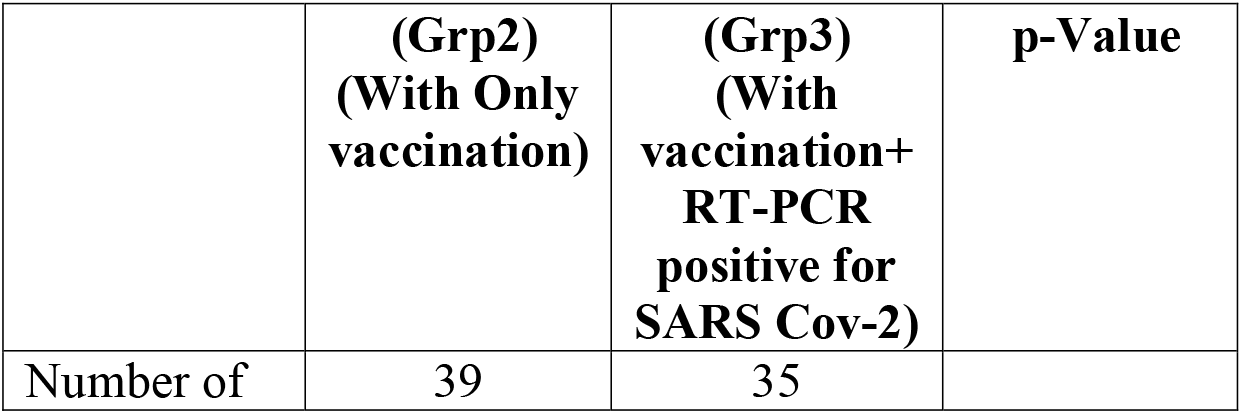
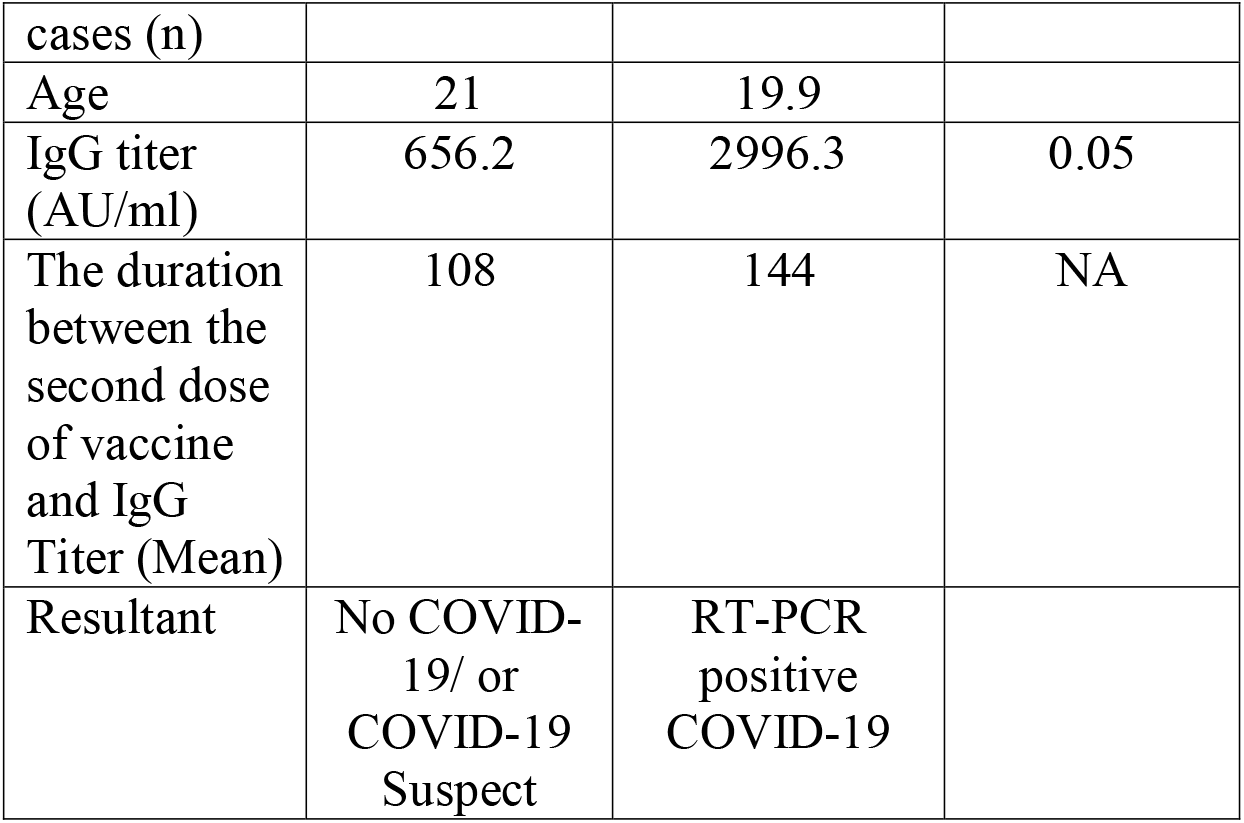
Showing Bolstering of IgG titer after vaccine+ Infection:

|  | **(Grp2)**<br>**(With Only**<br>**vaccination)** | **(Grp3)**<br>**(With**<br>**vaccination+**<br>**RT-PCR**<br>**positive for**<br>**SARS Cov-2)** | **p-Value** |
| --- | --- | --- | --- |
| Number of | 39 | 35 |  |
| cases (n) |  |  |  |
| Age | 21 | 19.9 |  |
| IgG titer (AU/ml) | 656.2 | 2996.3 | 0.05 |
| The duration between the second dose of vaccine and IgG Titer (Mean) | 108 | 144 | NA |
| Resultant | No COVID-19/ or COVID-19 Suspect | RT-PCR positive COVID-19 |  |

**Table 5:** Showing waning of IgG titer:

|  | **(Grp1)**<br>**IgG Titer**<br>**did after**<br>**60 Days** | **(Grp2)**<br>**IgG Titer**<br>**did after 150**<br>**Days** | **Significance** |
| --- | --- | --- | --- |
| Number of cases (n) | 31 | 39 |  |
| Age | 42.8 | 21.0 | p=0.000 |
| Time since vaccination | 60 | 150 |  |
| IgG titer Mean (AU/ml) | 1430.8 | 656.2 | p=0.034 |
| Resultant | No COVID-19 reported | No COVID-19 reported |  |

## Discussion

Breakthrough infections were operationally defined as the occurrence of COVID-19 infection ≥14 days after administration of two doses of COVID-19 vaccine irrespective of the type. In the present study, 35 experienced a breakthrough infection after being administered both scheduled doses of either COVID-19 vaccine.

Furthermore, nearly, 3 individuals reported the incidence of COVID-19 infection after receiving one dose of a COVID-19 vaccine but to before either complete vaccination or before 14 days post administration of the second vaccine dose. However, in our study, we also observed that of despite the infection, the disease in the infected was often mild or sometimes with nil symptomatology. Though very few similar studies on breakthrough infections are reported from India.^8,9^ Also, we observed that following a COVID-19 appropriate and COVID-19 preventive rules such as the use of face masks, maintaining social distancing, proper and regular hand washes, limiting social contact and gatherings, etc., had a huge impact on containing the transmission of infection among the exposed. It is beyond doubt that vaccines may have a greater impact on reducing viral circulation than naturally acquired immunity. The study by Tartof et al. observed that vaccines decrease disease severity among individuals who do acquire infection i.e., breakthrough infections. ^10^ The mechanisms by which COVID-19 vaccination elicits disease attenuation are largely unknown but are likely due to the recall of immunologic memory responses that reduce viral replication and accelerate the elimination of virally infected cells. Studies of viral dynamics further suggest that while viral loads in breakthrough infections may be as high in vaccinated individuals as they are in unvaccinated individuals, viral loads in those who are vaccinated decline more rapidly, and the virus that they shed is less likely to be culture-positive than virus shed by unvaccinated individuals.^11,12^ This suggests that people who are fully vaccinated are less likely to become infected and if infected, will be contagious for shorter periods than unvaccinated.^13^ Vaccines not only decrease transmission rates but also decrease disease severity among individuals who do acquire infection. This is very well supported by the present study by two means, one the breakthrough infection found in vaccinated individuals after an average lapse of 160 days was found to be very mild. So also, in the small group of SARS Cov-2 hospitalized patients, those who contracted an infection after an average of 30 days of vaccination, had no mortality despite their needed hospitalizations and oxygen therapy. (Table No: 4) To concerning delta variant, when vaccinated subjects with breakthrough infections are compared with unvaccinated, are less likely to develop mild or severe symptoms, more likely to recover from their illness quickly, and much less likely to require hospitalization. Nonetheless, the fact that vaccinated individuals can still become ill enough with COVID-19-19 to be hospitalized is understandably concerning. Even though vaccines are widely regarded as the most effective means of halting and terminating the COVID-19-19 outbreak, vaccine efficacy differs in the real world due to a variety of factors than reported.^14^ Hence, other preventive measures such as COVID-19 appropriate behavior which determines the degree of exposure to the virus also play a major role in preventing breakthrough infections. And our study strongly underscored the role of COVID-19 appropriate behavior and its additive effect on vaccine protection.^15^ A valuable new study by Tenforde and colleagues affirms the vaccine efficacy against hospitalization also depends on patients’ immune status, age, vaccine preparation, time since vaccination, and COVID-19 appropriate behavior among other parameters studied. The study further suggests that vaccines along with appropriate preventive measures are associated with a lower likelihood of infection and also transmission.^16^ Vaccination is the better effect when combined with better CAB, as all those who contracted COVID-19 breakthrough infections fared on the lower scale on CAB score in our study. This may have underlined the message that avoiding those contacts that are on the lower scale on CAB score (whose behavior is not in compliance with guidelines to prevent respiratory droplet infections) and avoiding close and crowded rooms is better for infection prevention. This warrants better awareness programs for propagating the importance of COVID-19 appropriate behavior. Of given the recent variants (omicron) and imminent variants of whatsoever nature, our study underlines the importance of vaccination and CAB.

Antibody response to SARS-Co-V 2: Evidence through multiple studies shows that protective antibody titers even though correlate well at the population level, are still elusive at the individual level^17^. A positive antibody test is currently recommended only to detect an individual with prior infection but cannot be used to assess the level of protection provided by the immune system in post-vaccinated individuals. However, a study by Bates et al. suggests that even though the breakthrough infections in patients were mild, there is a substantial increase of humoral immunity similar to the findings in our study.^18^ Additionally, in the wake of newer variants of SARS-CoV-2 emerging at a rather fast rate, the breakthrough sera have also demonstrated variant cross-neutralization in the vaccinated individuals. This creates a scope for a broadened protective immune response if the existing 2 dose series are further supplemented by variant boosters with antigenic inserts similar to the emerging variants and further prolong the protection as seen in a study conducted by Falsey et al.^19^ Naturally, antibody response to infection takes several days to a week and level decreases over time. Quantitative SARS-Co-V-2 anti-spike IgG response is varied by prior infection status, age, sex, type of vaccine & number of doses received. Post-vaccine antibody response is increased over 2-4 weeks after 1st dose of vaccination. Vaccine dose and following breakthrough infection cause a surge in IgG response as reported by this study.^20^

The decline in IgG titers is expected; but how these titers relate to protection from subsequent re-infections (Symptomatic or severe COVID-19 disease and death) remain elusive. An individual with a low or undetectable level of SARS-COV -2 antibodies have an increased risk of shedding infectious viruses.^21^ Effectiveness of vaccination and antibody-based -correlates of this protection are a potential strength of serological testing to identify those who are immune. Though this is a humongous and complex but a very novel aim, for which larger and complex studies are required. Studies with ‘Long longitudinal cohorts with baseline immunology’ are needed. This small study throws some insights for future studies.

This present outbreak could be because a new unknown COVID-19 variant that demands genomic sequencing and continuous surveillance. Unusual outbreaks like this and changing ecology of pandemic calls for deeper genomic and mechanistic studies.

The B.1.617.2 (Delta) variant of (SARS-CoV-2) was first identified in the state of Maharashtra, adjoining to the place of the present study in late 2020, and spread throughout India, outcompeting pre-existing lineages including B.1.617.1 (Kappa) and B.1.1.7 (Alpha). ^22,14^ This variant had higher replication and spike mediated entry and better host immune evasion. The latest variant omicron is allegedly milder because of its better availability to immune cells. The ongoing battle between the immune system and the natural selection of potential viral mutants is highlighted by the present study.

This study also demands real-time and representative studies from different geographies to understand the changing scene of emerging variants. Until now, Very few studies have been reported from India ^8,9^

Limitation of the study: This is not an active surveillance study, so claims made are of limited power, still similar reportage can be of useful for surveillance from peripheral and diverse sites to complement active surveillance.

## Conclusion

COVID-19-19 vaccination may as of now be considered as one of the front line forces to defeat the novel SARS-CoV-2 to prevent, manage and overcome the pandemic. The vaccine induces humoral response which is reflected in IgG response against spike protein and effectiveness of the vaccine is shown in the prevention of infection, the mildness of infection, and likely protection against mortality. And if vaccination is supported by COVID-19-appropriate behavior, it increases the chances of scoring over the novel coronavirus. Since India is also facing the emergence of new variants and a surge in COVID-19 cases, it makes the case stronger for following the practice of hand hygiene, wearing masks, and maintaining social distancing in public places. The two-pronged approach of vaccination and COVID-19-appropriate behavior would go a long way in fighting not only this wave but also future waves and variants of the pandemic. The use of serum anti-spike COVID-19 IgG is restricted to know the status of SARS-CoV-2 seroconversion.

## Supporting information

Case record Form

## Data Availability

All data produced in the present study are available upon reasonable request to the authors.

## Acknowledgement

**We sincerely acknowledge efforts of Mr. Dnyaneshwar Jadhav for running statistical tests and add value to the article**.

