## Supplementary material for "‘Anatomy of SARS-CoV-2 outbreak of ‘vaccinated’: An observational case-control study of COVID-19 breakthrough infections, COVID-19 appropriate behavior and anti-spike-IgG response as a correlate of protection in Medical college students at Rural Medical College, India": Case record Form

| **Table 1: Details of the Participant: (Case/ Control):::::::::::::::** | | |
| --- | --- | --- |
| Name: and Roll No |  | |
| Residential Room No: |  | ID: |
| Floor No: |  |  |
| Bld Name |  |  |

| **Table2: Vaccine Details:** |
| --- |
| **Name of the vaccine Taken** |
| First dose date |
| Second dose date |
| Any medicine taken after vaccine (0/1) |
| Time between vaccine and start of Illness |

| **Table 3: Dates of the Present Covid illness:** | | |
| --- | --- | --- |
| Illness Start Date |  | |
| Illness End Date |  | |
| RT-PCR date: and Name of the lab: |  | CT value (if Known): |
| Last travel before illness | Date: | |
|  | To Place: | |

| **Table 4: Self assessment clinical symptomatology of Covid on Scale of: 0 to 10:** | | | | | | | |
| --- | --- | --- | --- | --- | --- | --- | --- |
| **Symptom** | **Scale ( 0 to 10)** | **Symptom** | **Scale ( 0 to 10)** | **Symptom** | **Scale ( 0 to 10)** | **Symptom** | **Scale ( 0 to 10)** |
| Febrile Sense |  | Loss of smell |  | Chest pain |  | Muscle ache |  |
| Chill |  | Loss of taste |  | Chest discomfort |  | Arthralgia |  |
| Cough |  |  |  | Shortness of breatg |  |  |  |
| Sore throat |  | Nausea |  |  |  |  |  |
| Runny nose |  | Vomiting |  | Loss of appetite |  |  |  |
| Sputum |  | Abdominal pain |  | Malaise |  |  |  |
| Nasal stuffiness |  | Diarrohoea |  | General weakness |  |  |  |
|  |  | Constipation |  | Headache |  |  |  |

| **Table 6: Intervention/ Medication needed:** | | |
| --- | --- | --- |
| **Medicine/ Intervention** | Response( 0/1) | Days of requirement |
| Requirement of O2 |  |  |
| Remdesevir |  |  |
| Any other |  |  |

| **Table 5: Lab Investigations if any** | |
| --- | --- |
| **Lab Marker** | Value |
| CRP |  |
| CBC |  |
| D-Dimer |  |
| CT (0/1) |  |
| X-ray (0/1) |  |

| **Table 7: Behavioral habits:** | | | | | | |
| --- | --- | --- | --- | --- | --- | --- |
| Item | | Response( 0/1) | | Follow/up comment | | |
| Food habits  (Veg (0)/ Non Veg (1)) | |  | |  | | |
| **Table 8: Newly started Alternative/ recommended/ popular supplementations taken after the start of the pandemic:**  **(**These practices were not employed before the start of the pandemic) | | | | | | |
| **Item** | | | Response (0/1) | | Follow/up comment | |
| Zinc (0/1) | | |  | | Dose: Duration: | |
| Vit D (0/1) | | |  | | Dose: | |
| Inj Vit B12 | | |  | | Dose: | |
| Homeopathy: Arsenic album ( 0/1) | | |  | | Dose: | |
| Ashwagandha (0/1) | | |  | | Dose: | |
| Guduchi (0/1) | | |  | | Dose: | |
| Any other ayurvedic: (0/1) | | |  | | Dose: | |
| **Table 9: Physical Exercise Practices:** (irrespective of pandemic) | | | | | | |
| Item | | | | Response (0/1) | | Follow/up comment/  Scale of regularity (0-10) |
| Yoga/ Su –salutations (0/1) | | | |  | |  |
| Aerobic exercise (0/1)  ( jogging/ cycling/ swimming / oudoor Sports ) | | | |  | |  |
| Jal-neti: | | | |  | |  |
| Campus Bi-cycle Use | | | |  | |  |
| **Table 10: Major risk factor for Covid:** | | | | | |  |
|  | | | | Response | | Details |
| H/o any co morbidity/ major illness: (0/1) | | | |  | |  |
| H/o any close covid contact: (0/1) | | | |  | |  |
| H/o past covid infection(0/1) | | | |  | |  |
| Wt= Kg | Ht= (mt) | | | BMI: | |  |

| **Table 11: Covid appropriate behavior: (Before present covid-illness)** | |
| --- | --- |
| Item | Response on scale ( 0 to 10) |
| Wearing of mask |  |
| Hand sanitizer |  |
| Hand wash |  |
| Social distance maintenance |  |
| Following travel restrictions |  |

| **Table 12: Covid appropriate behavior: (After present covid-illness)** | |
| --- | --- |
| Item | Response on scale ( 0 to 10) |
| Wearing of mask |  |
| Hand sanitizer |  |
| Hand wash |  |
| Social distance maintenance |  |
| Following travel restrictions |  |
